# Recurring Spatiotemporal Patterns of COVID-19 in the United States

**DOI:** 10.1101/2021.11.23.21266775

**Authors:** Hawre Jalal, Kyueun Lee, Donald S. Burke

## Abstract

We analyzed the waxing and waning patterns (“surges”) of reported SARS-CoV-2 cases from January 1, 2020 through Oct 31, 2021 in all states and provinces (n = 93) in the USA, Mexico, and Canada, and across all counties (N = 3142) in the USA. A correlation matrix of the 576 × 576 daily case incidence rates in the 50 US states generates a distinctive “checkerboard” pattern showing that the epidemic has consisted of seven distinct internally coherent spatiotemporal wave patterns, four in the first year of the epidemic, and three thus far in the second year. Geoclustering of state case rate trajectories reveals three dominant co-varying spatial clusters of similar case rate trajectories, in the northeastern, southeastern and central/western regions of the USA. The spatiotemporal patterns of epidemic year 1 have thus far been repeated (p<.001) in epidemic year 2. The “checkerboard” pattern of the correlation matrix of case trajectories can be closely simulated as three sets of interacting sine waves with annual frequencies of 1:1:2 major cycles per year, corresponding to the northeastern, central/western, and southeastern state clusters. Case incidence patterns in Mexico and Canada have been similar to nearby regions in the southern US and the northern US, respectively. Time lapse videos allow visualization of the wave patterns. These highly structured geographical and temporal patterns, coupled with emerging evidence of annual repetition of these same patterns, show that SARS-CoV-2 case rates are driven at least in part by predictable seasonal factors.

**Significance Statement:** Local COVID-19 rates wax and wane. Often these epidemic changes are attributed to localized human behavioral factors. Our finding of highly structured continental scale spatiotemporal patterns that cross state and national boundaries, coupled with emerging evidence of annual repetition of these same patterns, shows that COVID-19 transmission is driven at least in part by seasonal factors. Other epidemic factors such as vaccine coverage rates, or emergence of new strains like the Delta variant of SARS-CoV-2 appear to modify, but not totally eclipse, these underlying seasonal patterns. COVID-19 seasonal transmission patterns are associated with, and may be driven by, seasonal weather patterns. Predictability of these patterns can provide opportunities for forecasting the epidemic and for guiding public health preparedness and control efforts.

## Introduction

The public health approach to local COVID-19 surges has often been reactive, with increased control measures following rather than preceding epidemic surges. For example, vaccination rates increased in the southern US states only after high case rates were experienced during the summer of 2021 (*1*). Prevention efforts may be more effective if implemented earlier, based on credible forecasts (*2*).

In earlier work, we showed that the incidence of COVID-19 in the United States has waxed and waned several times since the start of the epidemic, generating wave-like spatiotemporal patterns (*3, 4*). In this prior analyses we used data until May 3, 2021, and we predicted that a summer wave in 2021 similar to the 2020 summer wave may occur in the southern states (*3*). In the current manuscript, we extend the period of analysis through October 31, 2021, and we use methods to define spatiotemporal clustering of case time series. Elucidation and characterization of these patterns may aid in understanding the disease dynamics, enable forecasting of future surges, and facilitate improved targeting of public health resources.

## Results

Examining the time series of total US cases per day reveals wave-like patterns that are generated from the rise and fall of case incidence rates, as shown in **Fig. 1A**. However, this national-level display can mask important spatiotemporal and seasonal patterns because it aggregates data across disparate spatial units which may have varying local temporal patterns. In **Fig. 1B** we examine how cases covary across the 50 US states over time. Specifically, this plot examines correlations in case rate time series trajectories across states for each date against every other date. The color of each pixel represents the level of correlation between 1 (red-brown, exactly the same pattern of state case rates on the two dates) and –1 (blue, exactly the opposite pattern of state case rates on the two dates). **Fig. 1C** provides an example of the correlation behind one single pixel in this figure (of a total of 596^2 = 355,216 pixels) at the intersection between state case incidence rates Aug 1, 2021 on the x-axis vs. Aug 1, 2020 on the y-axis. This plot shows that the correlation of case rates across the 50 states between these two dates is 0.79, demonstrating very similar patterns in the case rate patterns between these two dates one year apart. Combining all possible correlations between all date pairs, creates the “checkerboard” pattern shown in **Fig. 1B**. The figure is dominated by alternating square shaped patches of high positive or negative correlation (p< .001). Note that these correlations measure the similarities of the relative magnitude of case rates across the 50 states from one day to another, but not the actual magnitude of the case rates. There are at least three regular patterns that emerge from this checkerboard. The first pattern is on the main diagonal that shows the distinct roughly square shaped boundaries of six waves that coincide roughly with the seasons, as well as the start of a seventh wave in fall 2021 (which we call Wave 6a, as explained below). Note that unlike **Fig.1A** that shows Wave 3 as a single large temporal wave from fall through winter (Wave 3), Wave 3 in **Fig. 1B** is clearly displayed as two distinct processes with different geographical patterns, which we designate here as Wave 3a and Wave 3b. Similarly, Waves 5 and the currently emerging Wave 6a are difficult to separate in **Fig. 1A** while their separation is revealed in **Fig. 1B**. The second main pattern is the remarkable similarity in the timing and geography of the waves observed in 2021 as compared to the year before, 2020. These can be seen as regions of high correlations located 12 months off the diagonal, and labelled as “1∼4” at the intersection of spring waves of 2020 and 2021, “2∼5” for the summer waves and most recently 3a∼6a for the fall waves, forming a “shadow diagonal” parallel to the main diagonal but displaced by one year. The third pattern is the high correlation regions between the summer 2020 and winter 2021 (2∼3b) waves and between the winter 2021 and summer 2021 (3b∼5) waves. These alternating regions of high and low correlations gives rise to annual patterns in the fall and bi-annual patterns in the winter and spring. **Fig. 1D** reveals the case trends by US regions. These three regions are defined using a spatially constrained clustering algorithm that identifies regions with similarity in case rate trends. The algorithm was able to identify 3 clusters, one in the northeast, one in the southeast and one in the central and western regions of the US (northeastern, 11 states with 71 million persons; southeastern, 8 states with 67 million persons; central/western with 31 states and 187 million persons; see map). Comparing the patterns in **Fig. 1D** and those in **Fig. 1B** reveals that Waves 1 and 4 occurred in the springs of 2020 and 2021, respectively and were concentrated in the northeastern states. Waves 2 and 5 occurred in the summers of 2020 and 2021, respectively and were concentrated in the southeastern states. Wave 3a was by far the largest and started in the northern region of the centra/western states in the fall of 2020. Although this display does not distinguish between wave 5 and 6a, it shows that while case rates declined sharply in the southeast, the decline has been much slower in the central/western region and lately cases have been slightly increasing in this region shown with the uptick of case rates at the end of the trend line. Furthermore, Wave 3b occurred in the winter 2021 and was concentrated in the northeastern and southeastern states. Overall, at least one region has been involved in each season since the epidemic has started, with patterns in 2021 repeating those in 2020.

**Fig. 1.**
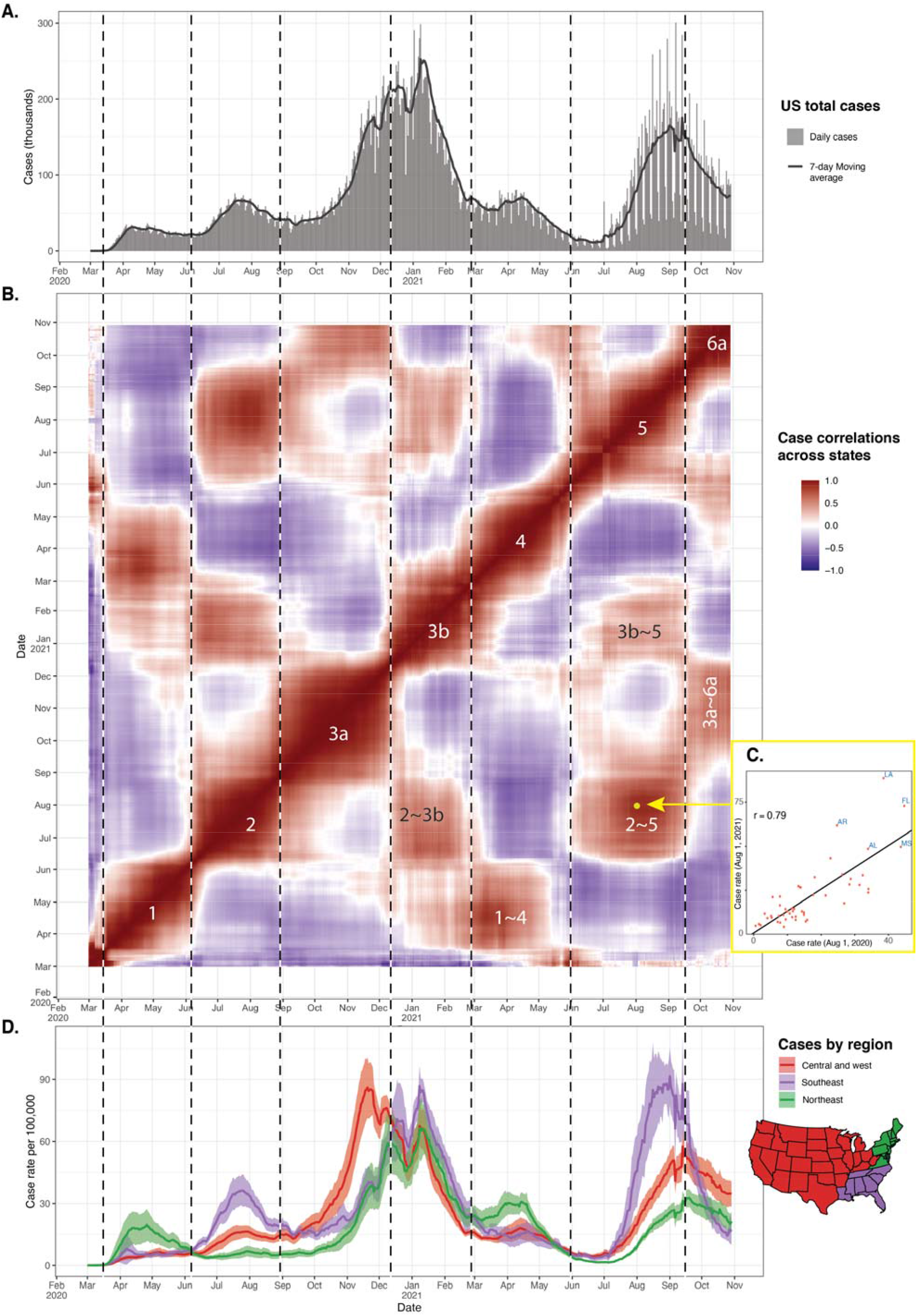
Correlation in daily COVID-19 case rates across states in the US from the start of the epidemic through Oct 31, 2021. Panel **A** shows the patterns of total cases and the 7-day moving average in the US. Panel **B** is a heatmap of correlations of daily cases across states. Each pixel in this figure represents the correlation in case rates between a date on the x-axis and another date on the y-axis. Panel **C** displays a scatterplot of case rates between August 1, 2020 and August 1, 2021 indicated by the yellow dot on Panel B (correlation r = 0.79). This scatterplot generates distinct regions of high and low correlations corresponding closely to the four seasons. The numbers on the diagonals indicate the wave indices: 1 = spring 2020, 2 = summer 2020, 3a = fall 2020, 3b = winter 2021, 4 = spring 2021, 5 = summer 2021, and 6a = fall 2021. The case rates in the spring, summer and fall seasonal waves in 2020 are highly correlated with the corresponding seasonal waves in and 2021. These regions are indicated with the labels 1∼4, 2∼5 and 3a∼6a in Panel B. In addition, Panel B shows at least two additional areas of high correlation that occur in different seasons (e.g., 2∼3b between summer 2020 and winter 2021, and 3b∼5 between winter and summer 2021). Panel **D** shows the average case rate across state by 3 contiguous regions defined with a spatially constrained clustering algorithm as described in the methods section. The states in each region are shown in the map under the legend. The vertical lines indicate the boundaries of each wave as recognized from Panel B. Notice that these boundaries do not necessarily coincide with visible wave patterns for the total US cases or cases by region especially for waves 4 through 6a because of overlapping wave patterns in Panels A and D.

Next we sought to simulate this specific “checkerboard” pattern using simple mathematical functions. The results are shown in **Fig. 2**. This figure is intended to simulate and reproduce the relationships between the overall case trends in **Fig. 1A** and those in the checkerboard in **Fig. 1B**. The deviation of the daily case rates from the average is shown on the top (**Fig. 2A**). In contrast to **Fig. 1D**, this figure shows the relative rise and fall of case rates compared to the US national average. Each deviation wave is mostly contained within a specific season of the year. For example, in spring of 2020, the northeast was above the average, while the southeast and central/western regions were below the average. This display shows clearly that while case rates declined in all three regions in the fall of 2021, the rate of decline was faster for the southeastern region compared to the other regions in **Fig. 1D**, resulting in an upward trend in the deviation from the mean for the central and western states, similar to fall of 2020.

**Fig. 2.**
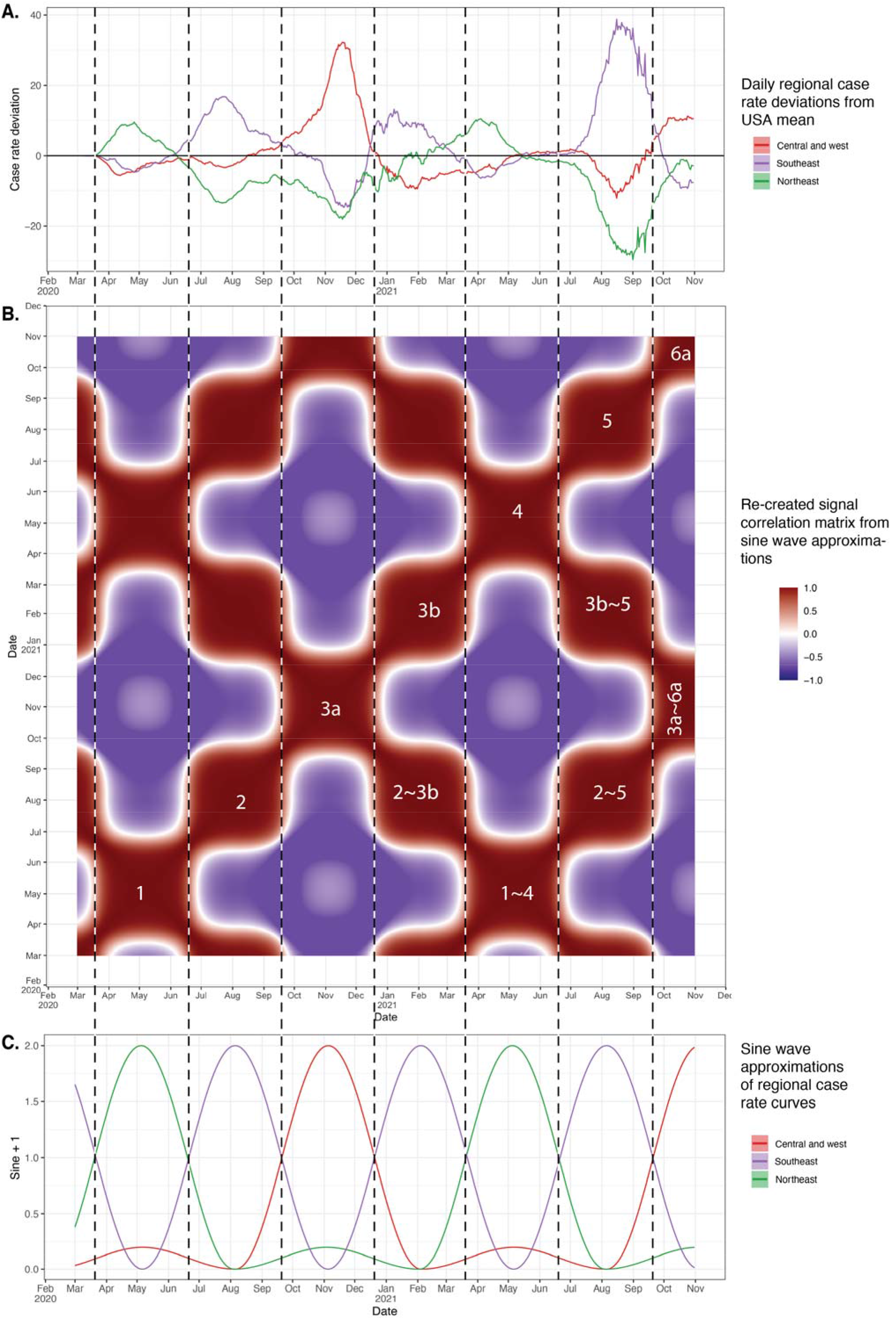
Reproducing seasonal patterns using a mixture of 3 sine waves. Panel A shows the deviation of daily case rates for the three regions from the daily average. Panel B presents a “checkerboard” pattern similar to Fig 1B above, but this version is based on a mixture of three simple sine waves – one per region as shown in Panel C. The three waves peak twice a year, but the waves for the central/western region and the northeast region alternate between a strong wave and a weak wave, whereas the amplitude of the southeast sine wave is fixed.

**Fig. 3.**
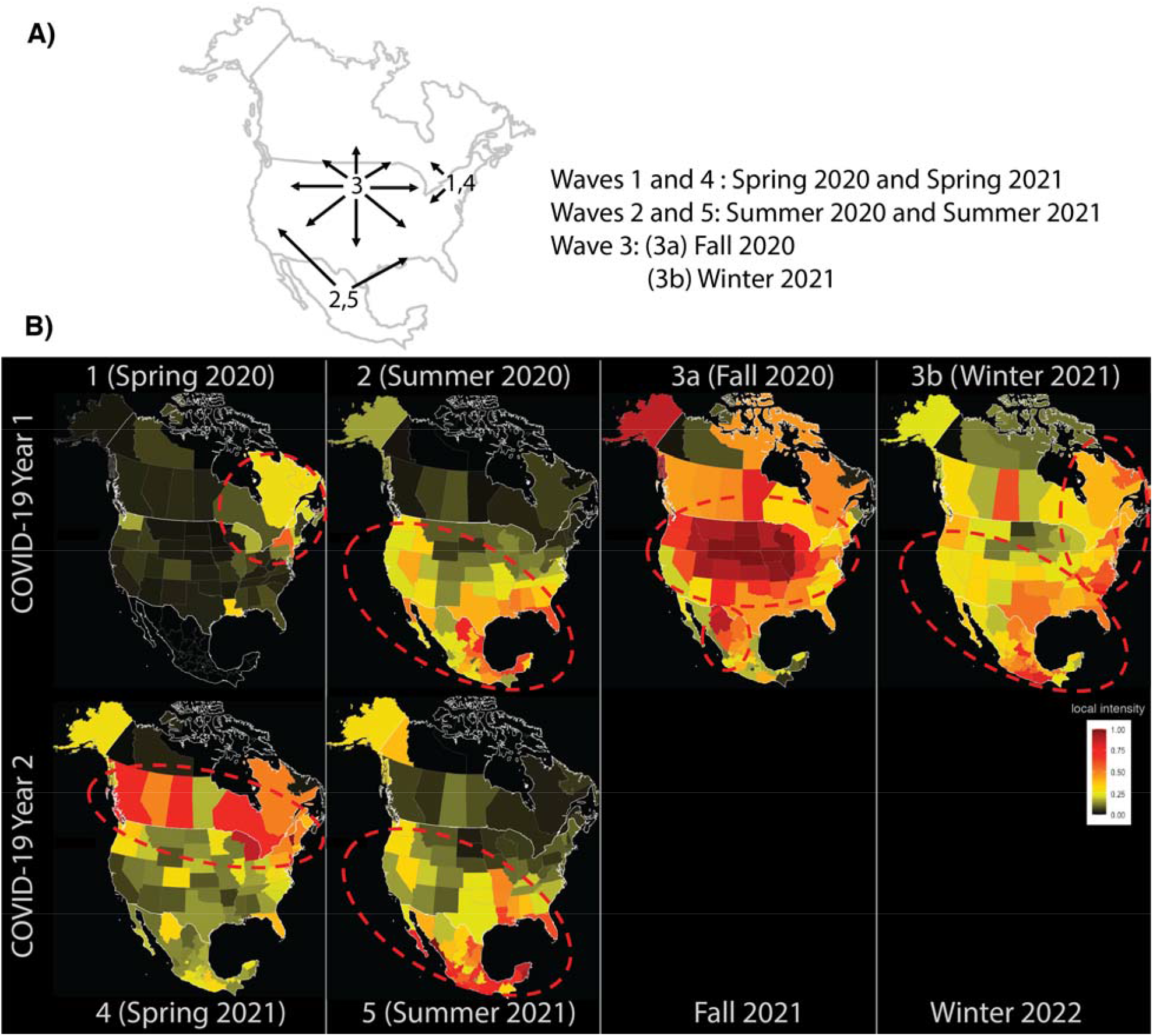
Spatiotemporal patterns of COVID-19 waves in the US, Mexico, and Canada by season. Panel **A** is a diagram illustrating the three major seasonal waves that has spanned the continent. The arrows indicate the direction of the spread. Panel **B** shows frames taken from **Movie 2** and reveals the geographical distribution of each wave and the similarity between the first and second year of the pandemic. The color scales represent the local intensity of cases on a scale from 0 to 1. As the movie shows, Wave 3a (fall 2020) continues through the winter months of 2021 (Wave 3b) as a travelling wave. The spring frames are for Apr 10, 2020 and 2021 (Waves 1 and 4), the summer frames are on July 28, 2020 and 2021, the fall frame represents Nov 20, 2020 (Wave 2 and 5), and the winter frame represents Feb 15, 2021 (Wave 3b). Mexico’s state level data for Wave 1 was missing.

We then approximated these dynamic patterns using three simple sine waves – one per each region shown in **Fig. 2C**. In this exercise, each sine wave peaks twice a year (wavelength = 6 months). However, the wave for the northeastern region and the central/western regions have alternating wave patterns in which they have a dominant peak in one season and a much smaller peak (10% of the full amplitude) six-months later. These waves result in a summer and winter wave for the southeastern region and a predominant fall-winter wave in the central/western region and the northeastern region. Examining the daily correlations among these three sine wave patterns produces a checkerboard pattern very similar to what we observe when examining the correlations among all 50 states (similarity of the sine wave simulation matrix heat map in **Fig. 2B** and the actual 50 state data matrix heat map in **Fig. 1B**).

The availability of US data at the county level provides an opportunity to further examining these seasonal waves. We created an animation that displays the changing spatiotemporal hotspots of COVID-19 case incidence by county using Getis-Ord Gi* statistic (**Movie 1**). The Gi* statistic represents the number of standard deviations above and below the national average for case incidence. There are several patterns that can be seen in this animation: First, the hotspots in each wave are defined to specific geographical region within seasons that repeat in the second year of the epidemic. For example, Waves 1 and 4 (springs waves) are mostly in the northeast, Wave 2 and 5 (summers waves) are in the southeast, while waves 3a and 6a (fall waves) are in the north central. Wave 3b which appears to propagate from 3a in winter of 2021 to the south has not repeated yet. Second, the hotspots in most of the waves spread to wide geographical areas beyond state boundaries despite variation in policies and biases in reporting, suggesting that the influence of large-scale factors, such as weather. Close observation of Wave 3 shows the gradual county-by-county movement of the epidemic from west to east from Iowa through Illinois, Indiana, Ohio, and Pennsylvania to New York and New England over the period of October 15 through December 15, 2020.

Next, we studied the relationship of the spatiotemporal patterns in US states to those across the border in Canada and Mexico using an animation of daily local intensity scores in COVID-19 incidence by states and provinces in the US, Canada, and Mexico (**Movie 2**). **Fig. 2** takes six frames from this video: one frame per wave summarizing the patterns described in this movie. Wave 1 (spring 2020) started in New England and Eastern Canada, with subsequent scattered intensity in the northern half of North America. Wave 2 (summer 2020) displayed a travelling wave pattern, starting from Mexico and southern and western states in the US. It first appeared in Mexico, crested northward across the border to the southern and western US states, then ebbed back southward. Wave 3a and 3b (fall 2020 and winter 2021) appear as a single wave in this presentation. Wave 3 emerged as a travelling wave from the Dakotas, flowing outward in a radial pattern to essentially all of North America. The ebb of this wave followed along the same general geographical pattern as its flow, again starting in the Dakota region, and ebbing outward in a radial pattern across the whole continent. Wave 4 (spring 2021) emerged from the tail end of Wave 3 in New England and rose quickly across all of Canada and some northern USA states. Wave 5 (summer) has started from Mexico and traveled upward towards the southern states in a fashion similar to Wave 2. Wave 6a has also started in the north-central region but have not peaked at the time of this writing.

Since the results of our analysis point to the influence of large-scale factors, we explored the relationship between temperature as a weather variable and case incidence in the US. We found a strong correlation between the timing between the Wave 3 peak date in each state and the timing of state temperature changes. The details of the analyses and the results are presented in the Supplementary Materials (**Figs. S1-S3** and **Movie S1**). Further research is needed to examine the link between weather variables and cases.

## Discussion

We used daily case rates from the beginning of the epidemic through October 31, 2021 to reveal COVID-19 seasonality patterns in the US, and similar patterns in Canada and Mexico. We showed that COVID-19 case rates in the US have waxed and waned six times, and that each wave has had a specific seasonal spatiotemporal pattern. In the northeastern US, COVID-19 waxed and waned during the spring season in both 2020 (Wave 1) and 2021 (Wave 4). The 2020 summertime wave in the southern US (Wave 2) repeated in the summer of 2021. Currently, Wave 6a (fall 2021) is repeating the patterns of Wave 3a in fall 2020. Interestingly, the winter 2021 wave in the southern states (Wave 3b) recapitulated the geographical pattern of Wave 2, with the net effect being that the southern states have experienced local waves at six months, rather than annual intervals. These waves had considerable seasonal spatiotemporal structure across international boundaries, such that patterns seen in Mexico are similar to patterns in the southern US, and patterns in Canada are similar to the northern US states.

The repeating waves in all three countries at approximately the same time of the year, the striking geographical structure of the COVID-19 epidemic in these countries, and the obvious shifting north / south latitudinal gradients, taken together strongly suggest a role for seasonal weather-related factors as major drivers of the epidemic. The overall wave structure suggests a bimodal relation between weather and cases in the southern states. Although complex, such a pattern has been seen in other studies (*5*). However, establishing a direct causal link between weather and COVID-19 incidence is challenging. Many infectious diseases exhibit complex seasonal patterns that are believed to be multifactorial and involve multiple complex interactions (*6*). Environmental factors and human behaviors that repeat around the same time every year might influence COVID-19 seasonality like other respiratory viruses (*3, 7, 8*). COVID-19’s viral survival, and transmission rate might be affected by temperature and humidity (*9-13*). An association between COVID-19 and relative or absolute humidity has also been described (*11*). These environmental conditions may determine how long the virus persists in aerosols and on surfaces (*14*). However, many of these factors (e.g., temperature and absolute humidity) are strongly correlated, complicating causal inference (*15*). In addition, changes in human behaviors have been associated with changes in environmental factors such as increased indoor contact during the winter months (*16, 17*). It appears likely that different seasonal-associated mechanisms may account for COVID-19 seasonality, but determining the exact causal pathways warrants further research.

The causes of transitory state and county level waxing or waning of COVID-19 case incidence rates have been the subject of prior analyses. Some studies support the effect of social-distancing measures such as closing schools and stay-at-home orders and other non-pharmaceutical interventions such as mask wearing on reducing the growth rate of infections (*18-22*). No doubt these measures among other factors can affect the state-by-state intensity patterns, including travel, population density and demographics, viral variants, vaccine coverage, and other important factors. These factors might have impacted the severity of the waves (amplitude) but may have been less influential in determining the timing of these waves. A proper causal analysis for the COVID-19 waves should include all these factors and their interactions.

Whatever the exact causal mechanism, so far, the seasonality of COVID-19 in the US has been repeated with considerable regularity: Summer waves in the south, spring waves in the northeast, and fall waves appear to be currently repeating in the northcentral region. It seems highly likely that the fall wave will continue similar to previous fall, given its strong correlation with seasons and its geographical pattern in 2020. If the population remains at immunity levels that are lower than those required for herd immunity, and with the emergence of stronger variants, the seasonal forces that have driven the epidemic could result in measurable epidemic surges at the same times and same regions into the future.

COVID-19 seasonality and its repeating spatiotemporal patterns can be used as tools in the effort of forecasting the epidemic and aid in prevention and control policies. Similar spatiotemporal analyses of other large scale COVID-19 data sets could be done around the world, linking the local epidemic patterns to local seasonal weather patterns. Results of such comparable international analyses could extend and support possible new epidemic forecasting methods based on weather. Consideration of the identified geographical patterns can be used to improve preparedness and response planning for future surges in COVID-19 which can disproportionately affect different regions. Like influenza, timely vaccination and control policies of COVID-19 can optimize resources, reduce economic burden, and effectively prevent future epidemics (*23-25*).

This study has several limitations. There are inconsistencies and biases in case reporting among states and provinces and across states. In addition, reported incidence is a function of the number of tests administered which can fluctuate over time. To reduce these testing and reporting biases, we scaled cases within each state and province to focus on the timing of the waves rather than their intensity. In addition, at the county level, the Gi* statistic incorporates the variability in a county and its neighbors, thus reducing the noise from any individual county. In addition, it is not yet possible to determine exactly how seasons have affected COVID-19 incidence. It is likely that human and virus factors interact differentially at various seasons shaping the epidemic’s waves. As future research investigates this complex relationship, we recommend that policies and decisions should seriously consider the seasonal patterns of COVID-19.

This analysis shows that patterns of waxing and waning of COVID-19 incidence at the state and county level are driven by continental-scale seasonal and geographical patterns. This in turn suggests that future state and county level COVID-19 surges should be at least in part predictable, and therefore preventable.

## Materials and Methods

### Data sources

Daily COVID-19 incidence rates for Canada, US, and Mexico were obtained from both The New York Times’ and John Hopkins University’s (*26*) COVID-19 data repositories. We used the full series from the start of the pandemic through October 31, 2021, which was the latest date for which data was available at the time of writing this manuscript. All our analyses use the 7-day moving average unless it is otherwise specified.

### Statistical Analyses

To examine periodicity in COVID-19 incidence in the USA, we measured the correlation in case rates between each date and every other date across states. This type of analysis is commonly used in signal processes to identify repeating patterns. We chose this correlation analysis over standard time trends because they can clearly disentangle periods of “synchrony” in states’ COVID-19 incidence rates. We produced a colored heatmap that highlights periods of high correlations on a single display. Spatiotemporal trends repeat when daily case rates between two isolated periods have high correlations across states. In addition, we used a spatially constrained clustering and regionalization algorithm to identify contiguous regions in the US that are similar in case incidence. Specifically, we used the implementation of the SKATER (Spatial ‘K’luster Analysis by Tree Edge Removal) algorithm (*27*) in the R package spdep. Briefly, the algorithm uses graph partitioning to efficiently divide the US into contiguous regions of states with similar COVID-19 case trend patterns. We conducted a sensitivity analysis in which we changed the number of clusters from 2 to 10. We chose 3 clusters as the additional clusters had only a state or two in some clusters.

COVID-19 incidence data is available for the US at the county level. To reveal how COVID-19 hotspots progressed over time, we animated a spatiotemporal hotspot progression using the Getis-Ord Gi* method (**Movie 1**). The Gi* method is widely used to show geographical clustering of hot (high) and cold (low) spots of noisy geographical data (*28*). The Gi* statistic is essentially a z-score which indicates the number of standard deviations for case rates in a county (and its neighbors) being above or below the national average. We used a modification of this method (*29*) to adjust the Gi* score over time.

To reveal the propagation of each COVID-19 wave in North America, we produced a time-lapse movie of daily case incidence at the state and province level in Canada, USA, and Mexico (**Movie 2**). Due to variations in reporting among states and provinces, we scaled COVID-19 incidence within each state and province to a range between 0 and 1, where 0 indicates no cases, and 1 indicates the highest number of cases in the state or province during the entire period. Thus, this video illustrates the timing of the waves rather than their relative intensity across states and provinces.

## Supporting information

Movie 1 Animation of COVID Incidence by US Counties

Movie 2 Animation of COVID Incidence in Canada, US, and Mexico

Supplemenary Movie S1 Animation of COVID Incidence and Temperature by US Counties

## Data Availability

All data produced are available online from the New York Times and John's Hopkins University from the sources identified in the manuscript.

## Acknowledgments

This research was supported by internal funding from the Graduate School of Public Health of the University of Pittsburgh.

## Movie captions

**Movie 1. Animations of changing COVID-19 hotspots in US counties**. The lower part of the graph shows the average trend for case rates across counties. The colors on the map indicate hot- and cold-spots of COVID-19 case incidence by county measured by the number of standard deviations above or below the national average across time. The animation uses 14 day moving average of case rates from March 29, 2020 through October 31, 2021. The lower plot shows the corresponding average case incidence rate across counties to provide a reference point in relation to the epidemic curve.

**Movie 2. Animation of the continental spread of COVID-19 in the US, Canada, and Mexico**. The animation starts from March 20, 2020 and ends on August 27, 2021. The color scale represents the local intensity score between 0 and 1 within each state or province. Zero indicating no cases in a state and 1 indicates the maximum number of cases for that state or province during the entire observation period. This animation shows the spatiotemporal patterns of the 6 waves in the US and the similarity of cases in the northeastern states with patterns in Canada and the southern states with patterns in Mexico. Wave 1 (Spring 2020) is centered in the northeast US and east Canada, Wave 2 (Summer 2020) appears as a travelling wave from Mexico to south US, Wave 3 appears also reveals a travelling wave pattern, with Wave 3a (Fall 2020) starts in the north-central US and continues through Wave 3b (Winter 2021) in south US. Wave 4 (Spring 2021) resets the cycle anew in the northeast US, and Wave 5 (Summer 2021) is centered in Mexico and then southern US.

## Supporting Information (SI)

### Supplementary Text

An exploratory analysis of the relationship between temperature and case incidence.

In this supplementary analysis we explored the relation between temperature as a weather variable and daily covid-cases. Daily weather stations’ temperature data readings were obtained from the National Oceanic and Atmospheric Administration (NOAA) (*30*). In addition, we obtained daily temperature on a 1-kilometer grid for the US from the Daymet Version 4 Monthly Latency Daily Surface Weather Data (*31*). All data used in this analysis are publicly available through the sources identified.

We chose the fall-winter 2020-21 wave to explore the relation between temperature and case incidences because the fall-winter wave was by far the most severe and widespread in the USA. We used the mcp (*32*) package in R to fit a change-point model and measure the peak incidence date. The supplementary **Fig. S1** shows the results of the change point analysis by state. The 95% Bayesian credible intervals were obtained from the posterior distribution of these change-points.

Representing when a state’s temperature reaches a specific threshold is challenging. Temperature varies every day between a low and a high value and a state may reach that temperature a few times during a season. In our calculations, we used the 14-day moving average to smooth the temperature trends and focused on the first time when each state reached a specified threshold. We estimated the date on which each state reached a temperature threshold. We used daily average temperature from a one-squared kilometer grid raster of the US. Considering the variation in the distribution of population within each state, we weighted the temperature by the proportion of the state population living in each squared kilometer in that state (*33*). Then, we computed the 7-day moving average to smooth the temperature trends and measured the dates on which that state’s temperature curve reaches four thresholds (15, 10, 5, and 0°C) within +/-1°C. The supplementary **Fig. S2** illustrates the temperature trend for each state indicating date on which each state’s temperature reached 10°C as an example. ‘‘

**Fig. S3** reveals the correlation between the day on which a state reached a specific temperature and the day on which temperature peaked at that state. The correlation coefficients were 0.62, 0.71, 0.77, and 0.73 for temperature thresholds of 15°C, 10°C, 5°C and 0°C, respectively. These are strong indicators supporting the association between air temperature and COVID-19 cases in Wave 3. In addition, the **supplementary Movie S1**, compares the spatiotemporal patterns of case incidence and temperature. This supplementary movie shows that Waves 2 and 5 coincide with temperature peaking in the southeastern states in the summers of 2020 and 2021, and that the travelling Wave 3a and 6a coincides with the movement of the cold front in the 2020 and 2021 fall seasons.

**Fig. S1.**
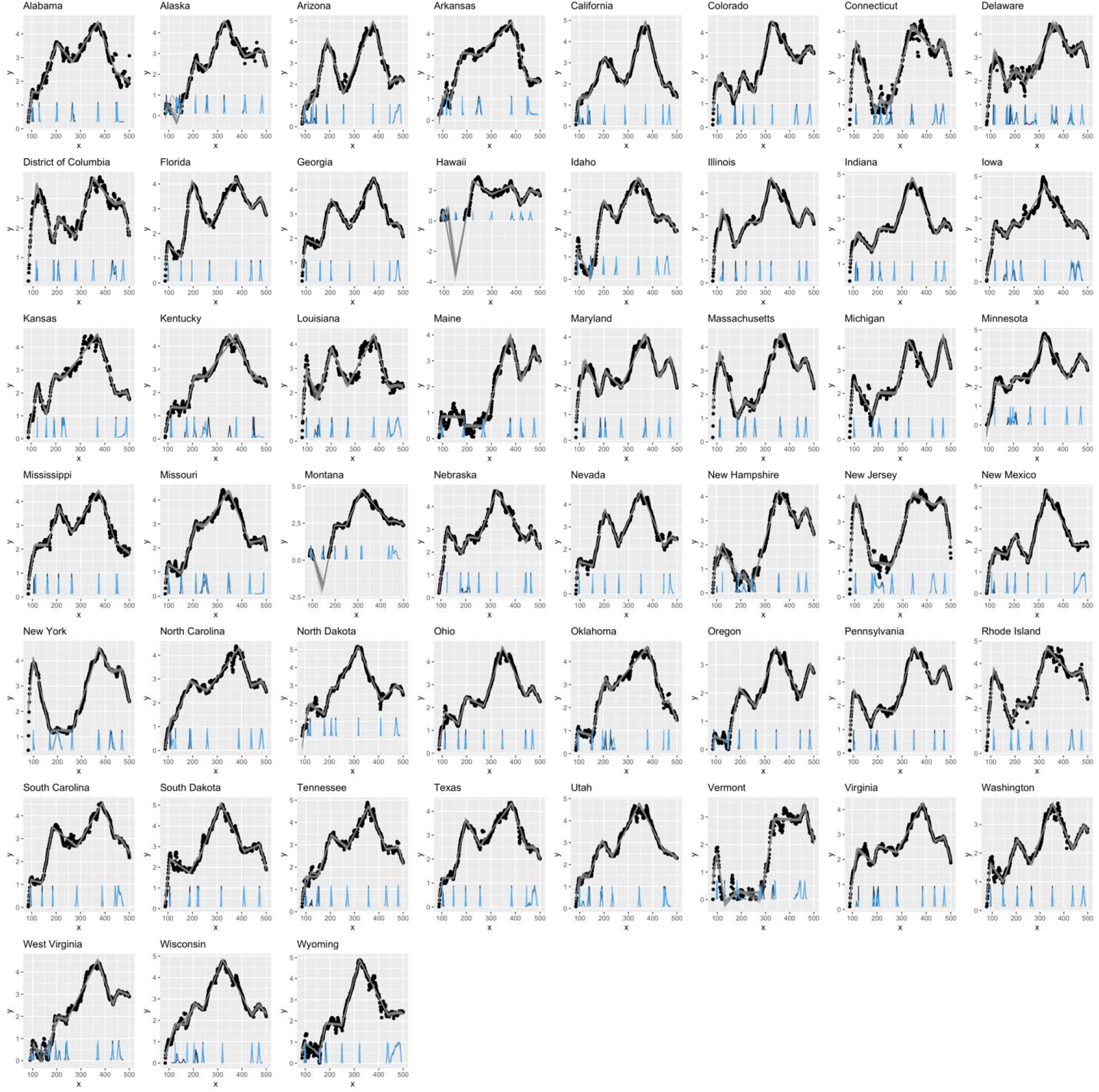
Results of the multiple change point analysis by state. The x-axis is the days since January 1^st^, 2020, and the y-axis is the log of 7-day moving average of case rates per 100,000. The gray lines are samples from the Bayesian posterior fitted piece-wise linear models, and the density plots at the bottom are the posterior densities of the seven change points. We used change points 1, 3, 5 and 7 to represent the days in which waves 1 through 4 peak, respectively.

**Fig. S2.**
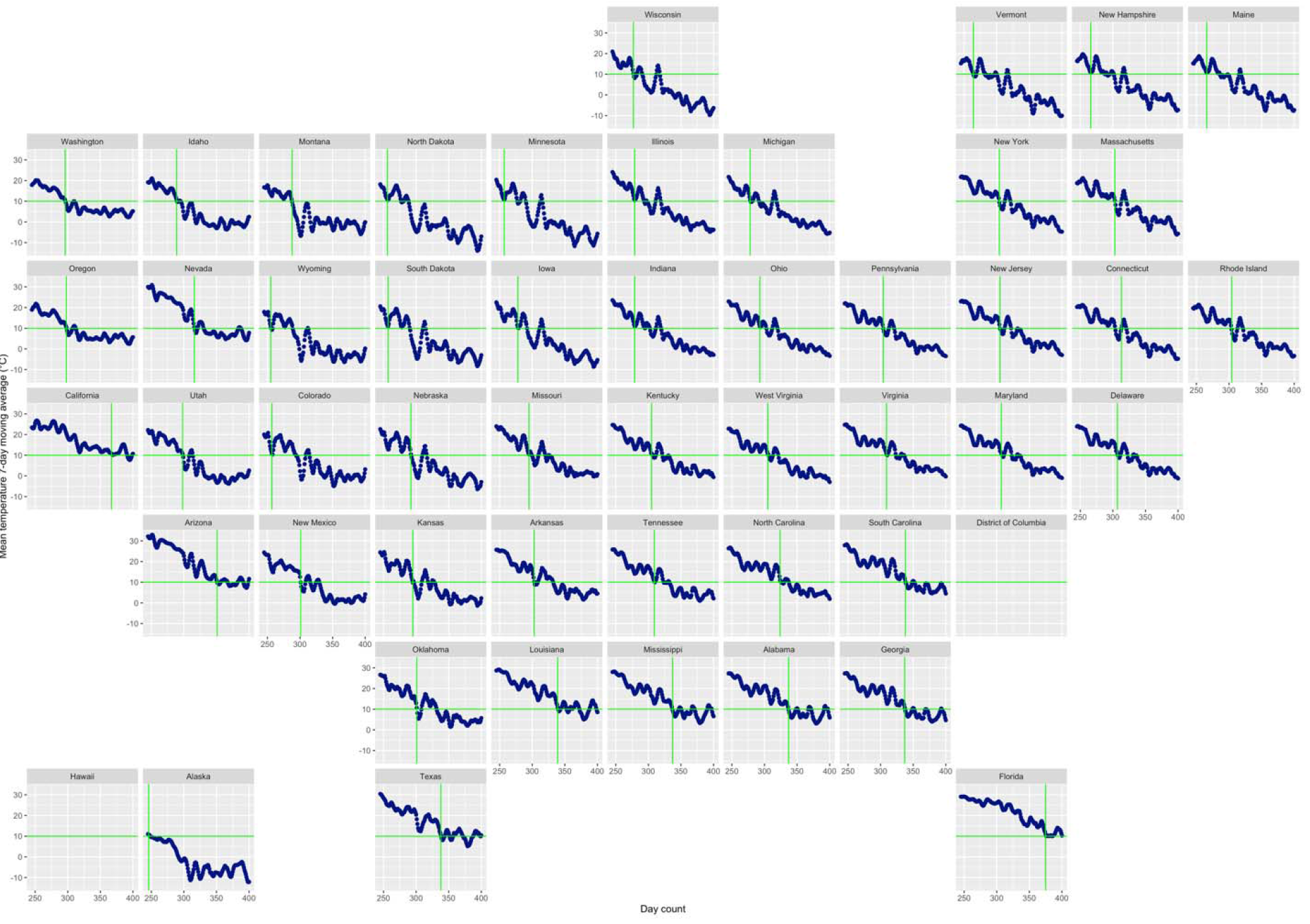
Measuring the date when temperature reaches 10°C. The x-axis is day counts since January 1^st^ 2020, and the y-axis is the mean temperature 7-day moving average. Each figure represents a state which are organized roughly by their locations in the US geological map. Data for the District of Columbia and Hawaii were not available. The horizontal green line indicates the temperature threshold used in this example (10°C), and the vertical line indicates the day at which the state’s curve reaches that threshold within +/-1°C.

**Fig. S3.**
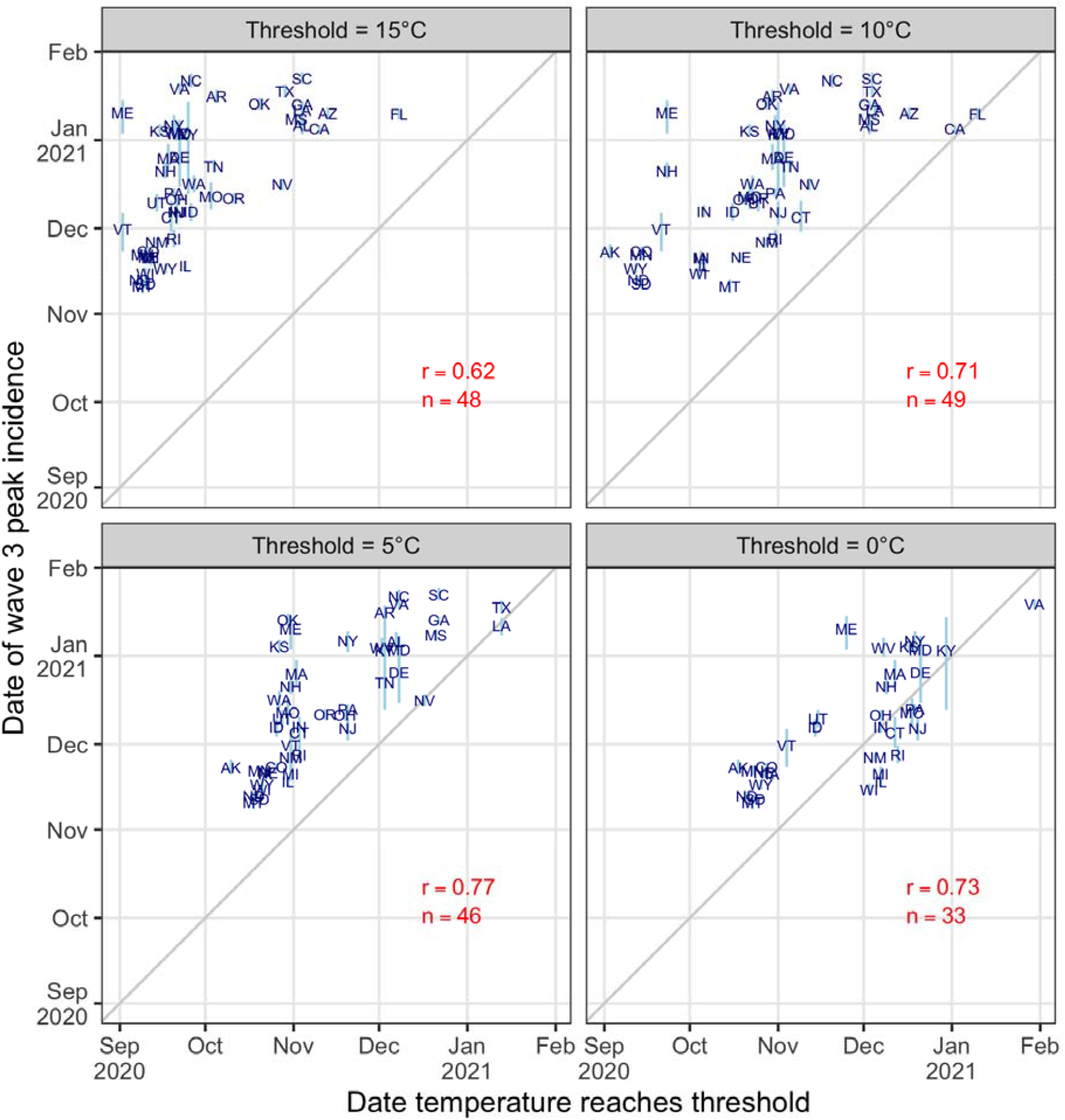
Temporal association between average temperature and Wave 3 peak incidence. Each plot shows a specific temperature threshold. The x-axis displays the date on which the average temperature in each state reaches the specified threshold within +/-1°C, and the y-axis is Wave 3’s peak incidence date. The 2-letter abbreviations represent the USA states. The light blue vertical lines are the 95% credible intervals for the peak dates, *r* is the correlation between the two dates across states, and *n* is the number of states that reach the temperature threshold. This analysis excludes Hawaii due to missing temperature data.

**Movie S1. Animations of average temperature and COVID-19 hotspots by county in the United States**. This movie shows the changes in temperature (left) and cases hotspots (right) as an exploratory display of the relationship. The lower part of the graph shows the average trend for both measures. The colors in the map on the left indicates average temperatures in degrees Celsius by county. The data was obtained by mapping the closest NOAA station to the centroid of each county. The colors for the map on the right indicate hot- and cold-spots of COVID-19 case incidence by county measured by the number of standard deviations above or below the national average (same as Movie 1 in the main text). The animation is from Jan 14, 2020 through October 31, 2021. Comparing the two animations reveals the spatiotemporal correlation between high temperatures in the southern states and the summer waves of 2020 and 2021, and the cooler temperature in the fall winter waves of 2020-21. In addition, the county-level visualization reveals additional details for spread of COVID-19 in the US, especially for Wave 3a and more recently Wave 6a.

